# Digital morphometry of tumor nuclei correlates to BAP-1 status, monosomy 3, gene expression class and survival in uveal melanoma

**DOI:** 10.1101/19010215

**Authors:** Christina Herrspiegel, Thonnie Rose O. See, Pia R. Mendoza, Hans E. Grossniklaus, Gustav Stålhammar

**Author notes:** Corresponding author Gustav Stålhammar, M.D. Ph.D. FEBO, Oncology and Pathology service, St. Erik Eye Hospital, Department of Clinical Neuroscience, Karolinska Institutet, Polhemsgatan 50, 112 82, Stockholm, Sweden.

## Abstract

Cytologic features such as the shape and size of tumor cells can predict metastatic death in uveal melanoma and other cancers but suffer from poor reproducibility. In this study, we investigate the interobserver concordance of digital morphometry, and correlate the results with BRCA associated protein-1 (BAP-1) expression and *BAP-1* gene mutation status, monosomy 3, gene expression classifications and patient survival in uveal melanoma. The average number of cells analyzed in each of 107 tumors, was 1957 (SD 349). Mean time consumption was less than 2.5 minutes per tumor. Identical morphometric classification was obtained for ≥ 85 % of tumors in all twelve evaluated morphometric variables (κ 0.70–0.93). The mean nucleus area, nucleus perimeter, nucleus max caliper and nucleus to cell area ratio were significantly greater in tumors with low BAP-1 expression and gene expression class 2. Patients had significantly shorter survival if their tumors had low BAP-1 (Log-Rank p=0.002), gene expression class 2 (p=0.004), long nucleus perimeters (p=0.031), long nucleus max calipers (p=0.029) and high mean nucleus to cell area ratios (p=0.041) as defined in a training cohort and then tested in a validation cohort. In the validation cohort, long nucleus perimeters and long nucleus max calipers correlated with monosomy 3 (Pearson Chi-Square p=0.006 and p=0.009, respectively). Long nucleus perimeters also correlated with *BAP-1* mutation (p=0.017). We conclude that digital morphometry can be fast and highly reproducible, that for the first time, morphometry parameters can be objectively quantitated in thousands of cells at a time in sub-μm resolutions, and that variables describing the shape and size tumor nuclei correlate to BAP-1 status, monosomy 3, gene expression class as well as patient survival.

## 1. Introduction

Uveal melanoma is the most common primary intraocular malignancy in adults (Singh *et al*., 2014). Less than 5 % of patients have clinically detectable metastases at the time of diagnosis (Singh *et al*., 2014). At a later stage however, up to 45 % of patients will develop metastases even if the eye containing the tumor has been removed (Kujala, Mäkitie and Kivelä, 2003). Once macrometastases develop, there is no effective treatment and median patient survival is only 4-12 months (Carvajal *et al*., 2016; Augsburger, Corrêa and Shaikh, 2009).

Several methods for prognostication are in clinical use. Tumor thickness, diameter, location in the eye and presence of distant metastases determine tumor stage (Kivelä *et al*., 2017; Arnljots *et al*., 2018). Loss of chromosome 3 has a high positive and negative predictive value for metastasis (Bornfeld *et al*., 1996). Commercial gene tests based on the expression of 12 classifier genes have been developed and show excellent prognostic utility in separation of class 1 tumors with low metastatic risk from class 2 tumors with high metastatic risk (Onken *et al*., 2012). Furthermore, we have previously shown the prognostic utility of manual (Szalai *et al*., 2018) and digital image analysis-based (Stålhammar *et al*., 2019a) determination of the level of nuclear BAP-1 (nBAP-1) expression.

In 1931, Callender described six types of uveal melanoma based on cytologic features such as cell shape and the size of the nucleus (Callender, 1931). The original classification could accurately predict metastatic death, but suffered from substantial intra- and interobserver discordance (Gamel, McCurdy and McLean, 1992; Coleman *et al*., 1996). After several modifications, the morphological classification of uveal melanoma now rely on assessments of the proportion of epitheloid tumor cells (McLean *et al*., 1983; Seddon *et al*., 1987). Examination of cytological features still require a high level of cytologic expertise and suffer from poor reproducibility (Gamel, McCurdy and McLean, 1992). Computer-assisted methods have therefore been proposed as a way of facilitating these assessments. In 1982, Gamel *et al*. found that 13 of 18 nuclear and nucleolar features correlated significantly with patient mortality when evaluated with a digitizer superimposed on microscopic images at a rate of 100 cells per hour (Gamel *et al*., 1982). Since then, computers have improved manyfold in terms of their computing power, cost and the number and scope of software applications and we can now analyze a dozen of variables or more in thousands of cells per minute on inexpensive off-the-shelf laptop computers (Stålhammar *et al*., 2018; Stålhammar *et al*., 2016).

Consequently, we see an opportunity to analyze cell morphometry features with digital image analysis and compare these to other prognostic factors including nBAP-1 expression in uveal melanoma patients from one American and one European referral center.

## 2. Methods

### 2.1. Patients and Samples

The study adhered to the tenets of the Declaration of Helsinki. Methods were carried out in accordance with the relevant guidelines and regulations. The protocol for collection of specimens and data from St. Erik Eye Hospital, Stockholm, Sweden was approved by the regional ethical review board in Stockholm, and the protocol for collection of specimens and data from Emory Eye Center, Atlanta, GA, USA by the Emory Institutional Review Board.

Patients in the training cohort (*n*=27) were identified in the archives of the Oncology and Pathology service, St. Erik Eye Hospital and L.F. Montgomery Ophthalmic Pathology Laboratory, Emory Eye Center. Inclusion criteria were: 1) Enucleation performed before December 2017, 2) Histologically proven uveal melanoma, 3) paraffin block available, 4) gene expression classification available, 5) clinicopathological data available, including tumor thickness, diameter, location, T-category and cell type, 6) follow-up data available, 7) sufficient tissue for BAP-1 immunohistochemistry. Exclusion criteria were: 1) Prior history of plaque brachytherapy, proton beam irradiation and/or transpupillary thermotherapy (TTT), and 3) tumor fully necrotic or fully hemorrhagic. 27 patients met the criteria. Our follow-up data was confirmed and further extended in telephone interviews with patients or relatives. Informed consent was obtained from all participants.

In order to establish generalizable morphometry thresholds for prediction of prognosis, we also included patients from the Cancer Genome Atlas (TCGA), made available by the National Cancer Institute at the National Institutes of Health (*n*=80). These patients had undergone enucleation due to primary uveal melanoma from 2011 through 2013, without previous brachytherapy, proton beam irradiation or TTT. Digitally scanned diagnostic slides were downloaded along with data on overall survival from TCGA on which the thresholds established in the training cohort were tested. *BAP-1* mutation and monosomy 3 status was downloaded from the supplemental information to the publication by Robertson *et al* (Robertson *et al*., 2017). No protected health information was accessed or downloaded from TCGA.

### 2.2. Immunohistochemistry

The paraffin blocks were cut into 4 μm sections, pretreated in EDTA-buffer at pH 9.0 for 20 minutes and incubated with mouse monoclonal antibodies against BAP-1 (clone C-4, Santa Cruz Biotechnology, Dallas, Texas, USA) and a red chromogen, and finally counterstained with haematoxylin and rinsed with deionized water. The deparaffinization, pretreatment, primary staining, secondary staining and counterstaining steps were run in a Bond III automated IHC/ISH stainer (Leica, Wetzlar, Germany). Dilutions between 1:20 and 1:500 had been evaluated before selecting 1:40.

### 2.3. Digital image analysis

After sectioning and staining, all glass slides were digitally scanned to the .ndpi file format at ×400, using identical digital scanners at both institutions (Nano Zoomer 2.0 HT, Hamamatsu Photonics K.K., Hamamatsu, Japan). The digital image analysis (DIA) software used was the QuPath Bioimage analysis v. 0.2.0 m4 (Bankhead *et al*., 2017). The software was run on a standard off-the-shelf laptop computer (Apple Inc. Cupertino, CA).

For assessment of the level of nBAP-1 expression, one positive cell (red chromogen in nucleus) and one negative cell (haematoxylin but no red chromogen in nucleus) was calibrated in each digitally scanned tissue section. All other parameters were left at default in order to limit time consumption and maintain ease of use. Tumors were then screened under low magnification (40×) and the area exhibiting the most intense nBAP-1 staining selected for grading. Nuclear immunoreactivity was evaluated at 200×, in a circular 0.5 mm-diameter region of interest (corresponding to the field of view in a light microscope with a 400× objective) by automatic classification (positive cell detection). Based on previous publications, the nBAP-1 expression was classified as “high” if immunoreactivity was detected in >30 % of tumor cells within the region of interest, and “low” if it was detected in ≤ 30 % of tumor cells (Stålhammar *et al*., 2019a; Szalai *et al*., 2018; See *et al*., 2019).

A workflow for morphometric analysis was then created, including the following steps for each tumor: A) Identification of all cells within the same circular 0.5 mm-diameter region of interest used for assessment of the level of nBAP-1 expression, using the software’s cell detection function with the following settings: Background nucleus radius 8 μm, median filter radius 0 μm, sigma 1.5 μm, minimum nucleus area 7.5 μm^2^, maximum nucleus area 200 μm^2^, threshold 0.1, max background intensity 2 and cell expansion 5 μm. B) Measurement in each detected cell in each region of interest of the following 12 cell morphometric variables: 1) Nucleus area (μm^2^). 2) Nucleus perimeter (μm). 3) Nucleus circularity. 4) Nucleus max caliper (μm). 5) Nucleus min caliper (μm). 6) Nucleus eccentricity. 7) Cell area (μm^2^). 8) Cell perimeter (μm). 9) Cell circularity. 10) Cell max caliper. 11) Cell min caliper. 12) Nucleus to cell area ratio (figure 1).

**Figure 1.**
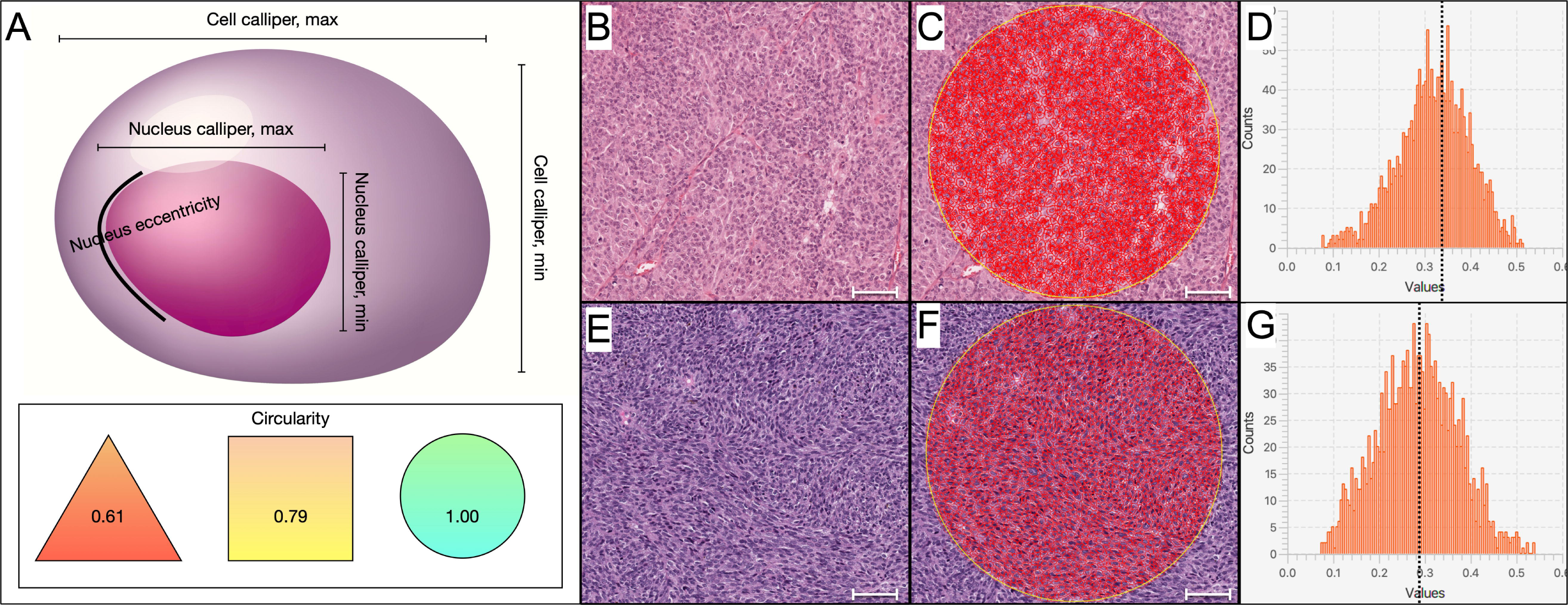
Illustration of cell morphometric measurements. A) Calipers denotes the largest and smallest diameters of the nucleus and cell. Nucleus eccentricity is a measure of how much the nucleus deviates from a spherical shape, presented as a number between 0.00 and 1.00. A completely spherical nucleus has an eccentricity of 0.00, a nucleus with the shape of an elliptical 3D solid would have an eccentricity of 0.5, whereas a 3D conical distribution would have a value of 1.00. Circularity compares the perimeter of a shape to the area it contains and is calculated by four times π times the area divided by the perimeter squared. The circularity of a circle is 1.00, and less for less circular objects. The nucleus to cell area ratio is the area of the nucleus divided with the area of the entire cell. B) Example of the morphologic appearance of a tumor mainly composed of epitheloid cells. C) In this tumor, a circular 0.5 mm-diameter region of interest (corresponding to the field of view in a light microscope with a 400× objective) has been identified. Within the region of interest, each cell is automatically analyzed for the level of nBAP-1 expression and 12 morphometric variables. D) The results of the measurements can be presented as frequency distributions. In this example, the mean nucleus to cell area ratio of the cells measured in the region of interest was 0.32 (dotted line). E) and F) A tumor mainly composed of spindle-like cells is shown for comparison. G) This tumor had a slightly lower mean nucleus to cell area ratio of 0.28. Cell illustration by iStock.com/Vitalii Dumma, East Ukraine Volodymyr Dahl National University Scale bars: 100 μm.

Tumor areas with intense inflammation, heavy pigmentation, bleeding, necrosis or poor fixation were avoided. nBAP-1 classification and morphometric analysis was performed blinded to all other patient data including outcome. For measurement of interobserver concordance, two human observers performed the digital morphometry (morphometric variable above or below median value) and nBAP-1 classification (high or low) independently and blinded to patient outcomes.

### 2.4. Gene expression classification

Tumor tissue samples were obtained from freshly enucleated eyes by fine needle aspiration. The contents of the needle hub were transferred into one of two RNAse-free cryovials. Using the same needle, extraction buffer from the second cryovial was aspirated and expelled into the first. This was then placed in a specimen bag, immediately frozen to -80° C and shipped on dry ice for gene expression classification based on 12 discriminating genes (*HTR2B, ECM1, RAB31, CDH1, FXR1, LTA4H, EIF1B, ID2, ROBO1, LMCD1, SATB1*, and *MTUS1*) and 3 control genes (*MRPS21, RBM23*, and *SAP130*) at a commercial laboratory (Castle Biosciences Inc. Friendswood, TX, USA). Expression levels of the gene products are used to categorize tumors as either class 1 with low metastatic risk, or class 2 with high metastatic risk (Onken *et al*., 2012).

### 2.5. Statistical methods

Differences with a p<0.05 were considered significant, all p-values being two-sided. The deviation of all clinicopathological variables from normal distribution was statistically significant, when evaluated by the Shapiro–Wilk test (p<0.05). For statistical tests of these variables, we therefore used the Mann-Whitney *U* test, which does not assume normally distributed data. The deviation of all morphometric variables from normal distribution was however not statistically significant (p>0.05), why we used one-way ANOVA with Bonferroni correction for these. For comparisons of categorical variables, two-by-two tables and Fisher’s exact test were used. For correlation to Cox Proportional Hazards for metastasis and Kaplan-Meier metastasis-free survival, patients were split into two groups based on 1) the median value of each morphometric variable, and 2) receiver operating characteristics (ROC) in the training cohort, with equal emphasis on sensitivity and specificity for the development of metastasis. The thresholds established in the training cohort were then tested in the validation cohort, with Kaplan-Meier survival analysis and in two-by-two tables for Pearson Chi-Square correlation with *BAP-1* mutation and monosomy 3. In evaluation of interobserver concordance, the percentage of identically classified cases and Cohen’s kappa statistics (κ) were computed (Cohen, 1960). Metastasis-free follow-up was defined as the time in months from enucleation to the last occasion patients without metastases was seen or in contact alive. All statistical analyses were performed using IBM SPSS statistics version 25 (Armonk, NY, USA).

## 3. Results

### 3.1. Descriptive statistics

The mean age at enucleation of patients in our training cohort was 66 years (SD 15). Of 27 patients, 15 were men and 12 women. 25 tumors originated in the choroid and 2 in the ciliary body. The cell type was mixed in 18 patients, spindle in 5 and epitheloid in 4. Mean tumor thickness was 8.6 mm (SD 3.7) and mean diameter 15.8 mm (SD 4.8). 12 tumors were of gene expression class 2 and 15 of class 1a or 1b. 14 tumors had low nBAP-1 expression and 13 high. Mean metastasis-free follow-up time was 47 months (SD 76). The validation cohort had similar features (Table 1).

**Table 1.**
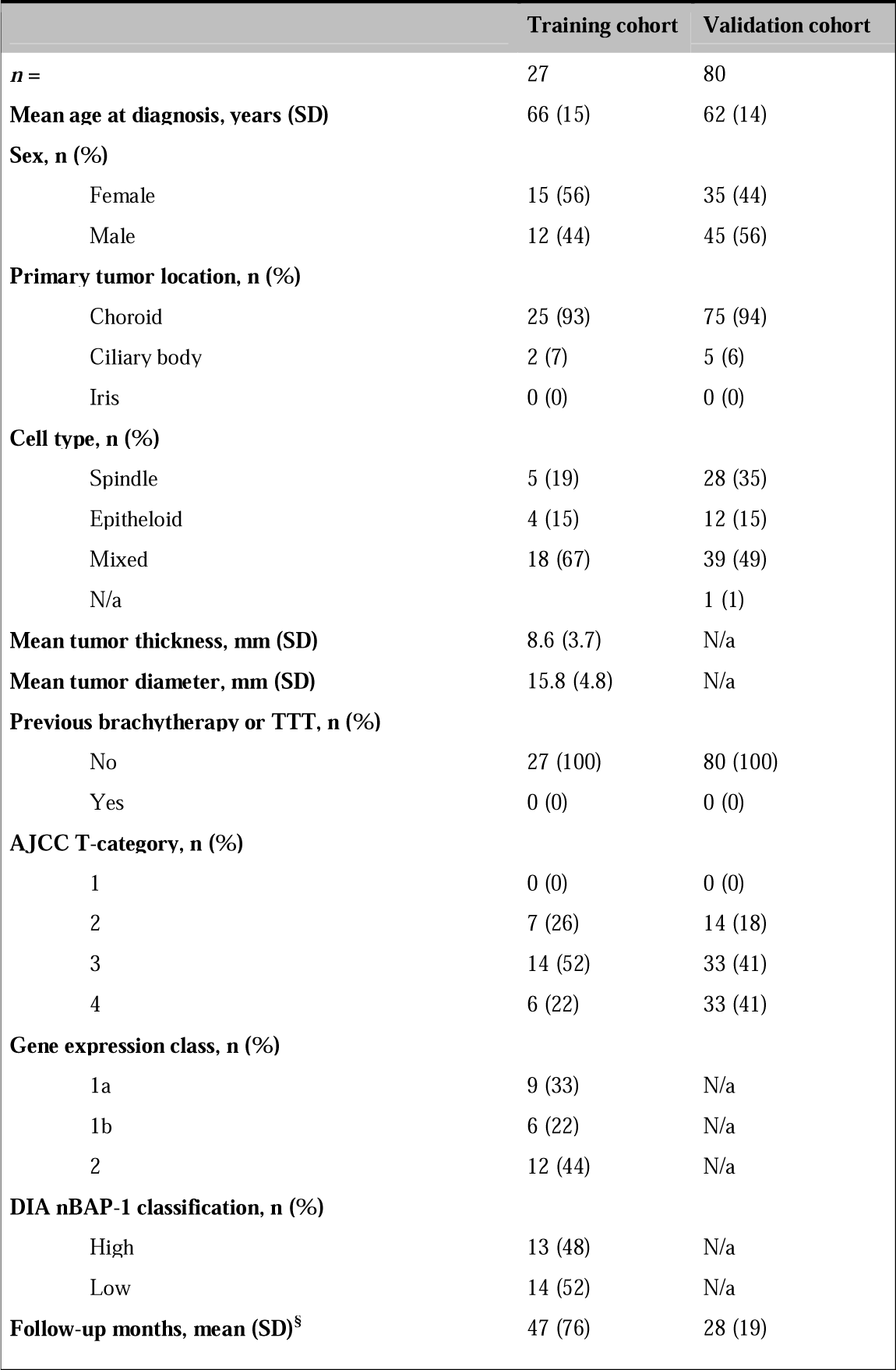
Characteristics of patients and tumors included in this study. SD, standard deviation. N/a, not available. TTT, Transpupillary thermotherapy.

The average number of cells analyzed in each tumor was 1957 (SD 349), which took an average of 74 seconds (SD 21) for nBAP-1 classification and 71 seconds (SD 17) for morphometric analysis, adding up to 145 seconds or nearly two- and-a-half minutes per tumor.

### 3.2. Interobserver concordance

Identical nBAP-1 classification was obtained for 25 of 27 tumors (93 %) in the training cohort, yielding a Cohen’s kappa statistic indicating almost perfect agreement (κ=0.85).

Identical morphometric classification (morphometric variable above/below median value) was obtained for ≥ 85 % of tumors in all 12 variables, yielding substantial or almost perfect agreement (κ 0.70–0.93, Table 2).

**Table 2.** Interobserver concordance and Cohen’s kappa statistics in classification of each morphometric variable as above or below the median value.

|  | **Interobserver concordance (%)** | **Cohen's $\kappa$** |
| --- | --- | --- |
| Nucleus area | 85 | 0.70 |
| Nucleus perimeter | 85 | 0.70 |
| Nucleus circularity | 96 | 0.93 |
| Nucleus caliper, max | 85 | 0.70 |
| Nucleus caliper, min | 89 | 0.78 |
| Nucleus eccentricity | 96 | 0.93 |
| Cell area | 85 | 0.70 |
| Cell perimeter | 85 | 0.70 |
| Cell circularity | 93 | 0.85 |
| Cell caliper, max | 85 | 0.70 |
| Cell caliper, min | 85 | 0.70 |
| Nucleus to cell area ratio | 96 | 0.93 |

### 3.3. Morphometry versus nBAP-1 expression and gene expression class

The mean nucleus area, nucleus perimeter, nucleus max caliper and nucleus to cell area ratio were significantly greater in tumors with low nBAP-1 expression. Nucleus circularity, nucleus min caliper, nucleus eccentricity and cell area, cell perimeter, cell circularity, cell max and min caliper were however not significantly different (Table 3a).

**Table 3.**
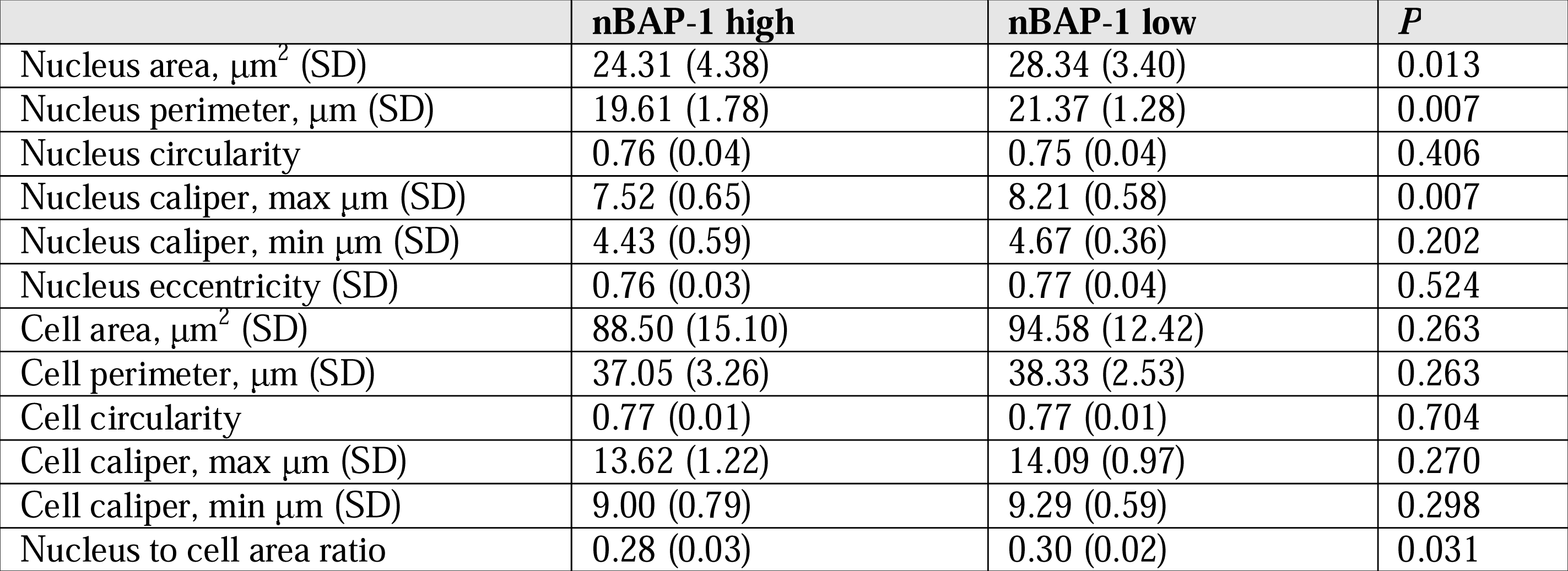
Average values and *P* defined by one-way ANOVA with Bonferroni correction of cell morphometric variables in tumors of high versus low nBAP-1 expression. SD, standard deviation.

Similarly, the nucleus to cell area ratio, but not the other morphometric variables, were significantly greater in tumors of gene expression class 2 (Table 3b).

**Table 3b.**
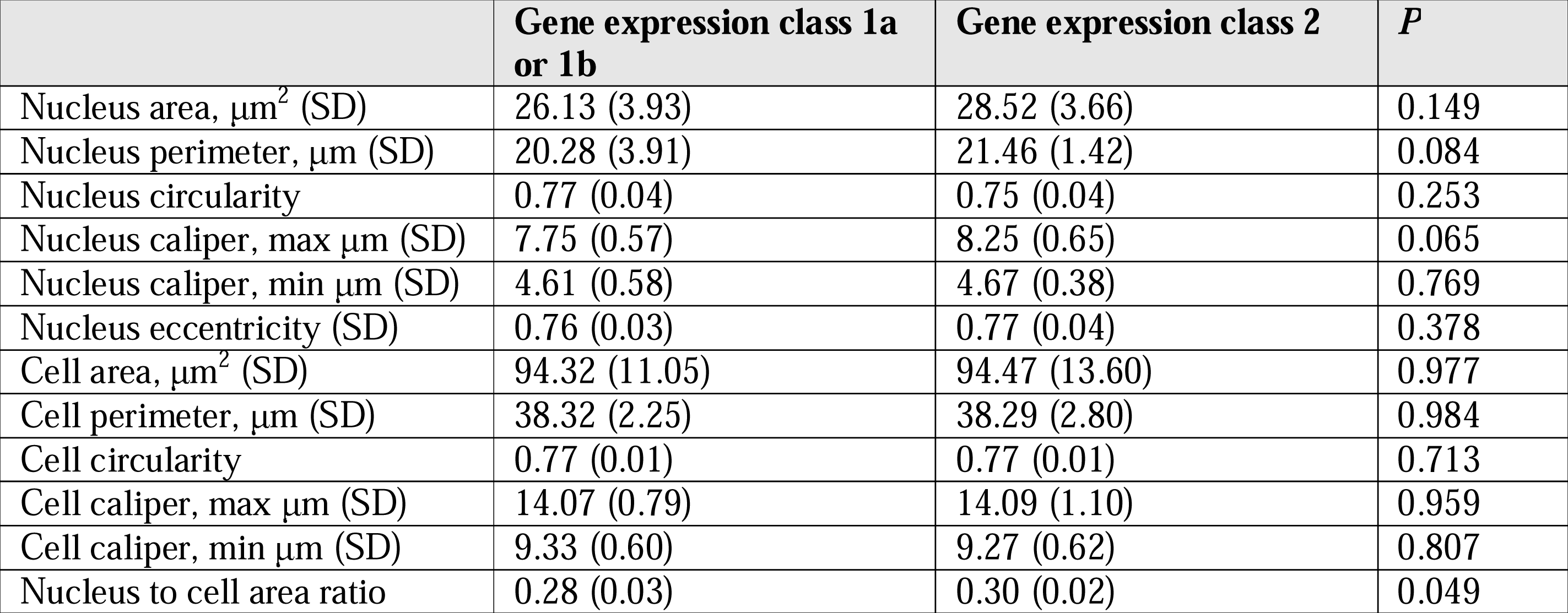
Average values and and *P* defined by one-way ANOVA with Bonferroni correction of cell morphometric variables in tumors of gene expression class 1a or 1b versus 2. SD, standard deviation.

### 3.4. Adjusted thresholds

The mean nucleus area, nucleus perimeter, nucleus max caliper and nucleus to cell area ratio were analyzed with ROC, with equal emphasis on sensitivity and specificity for the development of metastasis. Mean nucleus area achieved an area under the curve (AUC) of 0.54 (sensitivity 67 %, specificity 50 %, p=0.76) at threshold 28 μm^2^; Mean nucleus perimeter achieved an AUC of 0.58 (sensitivity 67 %, specificity 56 %, p=0.50) at threshold 22 μm; Mean nucleus max caliper achieved an AUC of 0.61 (sensitivity 56 %, specificity 67 %, p=0.38) at threshold 8.2 μm; and mean nucleus to cell area ratio achieved an AUC of 0.69 (sensitivity 56 %, specificity 67 %, p asymptotic significance p=0.12) at threshold 0.32 (figure 2).

**Figure 2.**
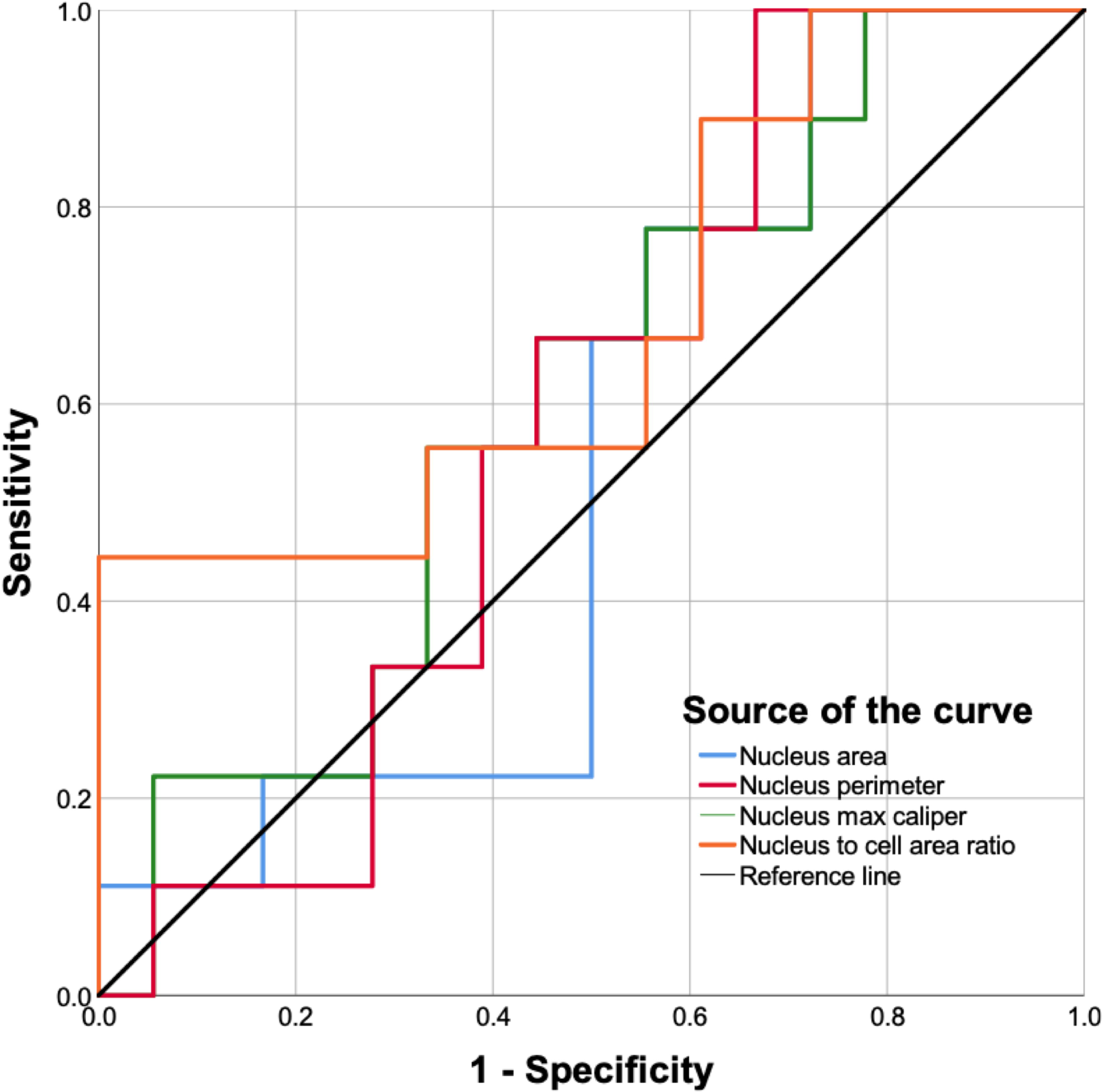
Receiver operating characteristics (ROC) of the mean nucleus area, mean nucleus perimeter, mean nucleus max caliper and mean nucleus to cell area ratio in the training cohort (*n*=27), with equal emphasis on sensitivity and specificity for the development of metastasis. Mean nucleus area (blue line) achieved an area under the curve (AUC) of 0.54 (sensitivity 67 %, specificity 50 %, p=0.76) at threshold 28 μm^2^; Mean nucleus perimeter (red line) achieved an AUC of 0.58 (sensitivity 67 %, specificity 56 %, p=0.50) at threshold 22 μm; Mean nucleus max caliper (green line) achieved an AUC of 0.61 (sensitivity 56 %, specificity 67 %, p=0.38) at threshold 8.2 μm; and mean nucleus to cell area ratio (orange line) achieved an AUC of 0.69 (sensitivity 56 %, specificity 67 %, p asymptotic significance p=0.12) at threshold 0.32.

### 3.5. Regression analysis and survival

In survival analysis, the thresholds defined in our training cohort was applied to morphometry measurements in the validation cohort. Kaplan-Meier overall survival was significantly shorter for patients with tumors that had long nucleus perimeters (Log-Rank p=0.031), long nucleus max calipers (Log-Rank p=0.029) and high mean nucleus to cell area ratios (Log-Rank p=0.041). Patients also had significantly shorter survival if their tumors had low nBAP-1 expression (Log-Rank p=0.002) or gene expression class 2 (Log-Rank p=0.004). The nucleus area was however not associated with shortened survival (Log-Rank p=0.266, figure 3).

**Figure 3.**
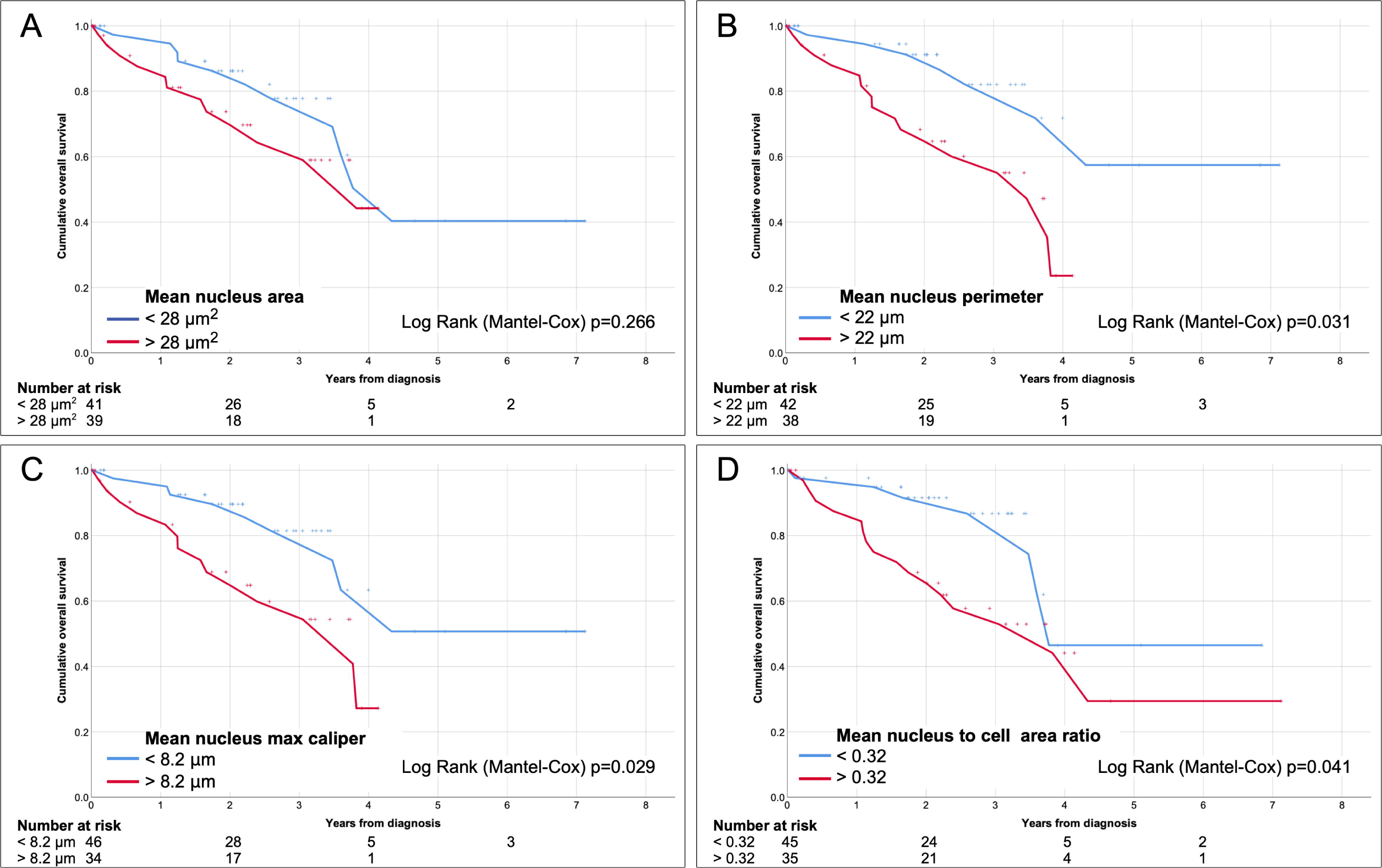
Kaplan-Meier curves, cumulative overall survival in the validation cohort (*n*=80). A) Patients with tumors with mean nucleus area < 28 μm^2^ (blue) versus > 28 μm^2^ (red, Log-Rank p=0.266). B) Patients with tumors with mean nucleus perimeter < 22 μm (blue) versus > 22 μm^2^ (red, Log-Rank p=0.031). C) Patients with tumors with mean nucleus max caliper < 8.2 μm (blue) versus > 8.2 μm^2^ (red, Log-Rank p=0.029). D) Patients with tumors with mean nucleus to cell area ratio < 0.32 (blue) versus > 0.32 (red, Log-Rank p=0.041). The thresholds were established in the training cohort.

In univariate Cox proportional hazards analyses of nucleus area, nucleus perimeter, nucleus max caliper and nucleus to cell area ratio, all variables were individual predictors of metastasis. In multivariate analysis, nucleus perimeter and nucleus to cell area ratio retained their significance (Table 4).

**Table 4.**
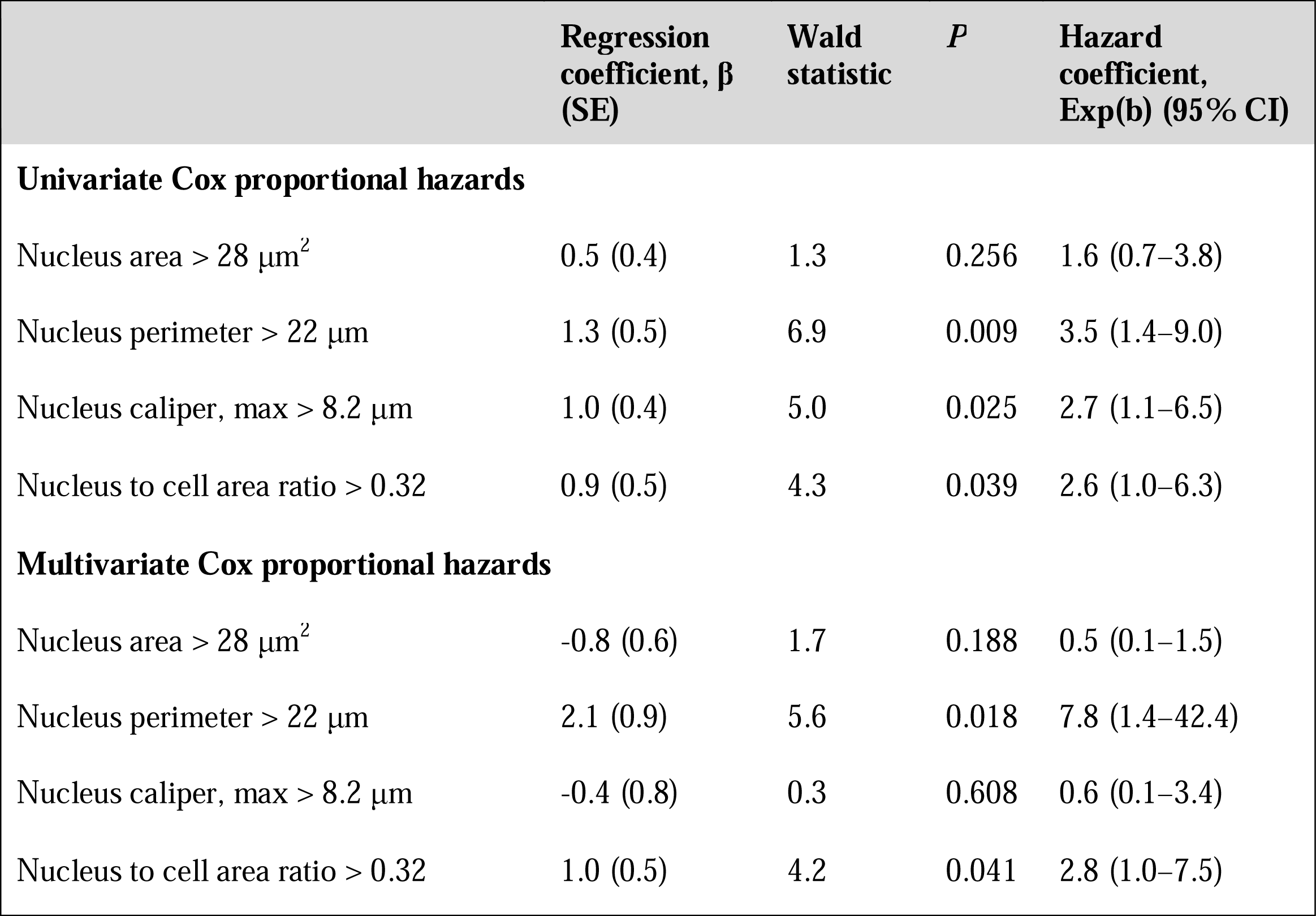
Univariate and multivariate Cox Proportional Hazards analysis of the association between metastasis-free survival and cell morphometric variables. SE, standard error.

In two-by-two tables, long nucleus perimeters correlated with *BAP-1* mutation and monosomy 3 (Pearson Chi-Square p=0.017 and p=0.006, respectively). Long nucleus max calipers correlated with monosomy 3 (p=0.009) but not with *BAP-1* mutation (p=0.085). Nucleus area and nucleus to cell area ratio did not correlate with *BAP-1* mutation or monosomy 3 (p>0.21).

## 4. Discussion

In this study, we have shown that digital morphometry of uveal melanoma can be fast and highly reproducible, and that variables describing the size of the tumor cell nuclei correlate to gene expression class, *BAP-1* status, monosomy 3 and patient survival. On the other hand, no variable describing the shape and size of the entire tumor cell correlated to the prognostic factors, indicating that for prognosis, the morphological characteristics of tumor nuclei are more important.

The prognostic importance of cell morphology is by no means a novel discovery or limited to uveal melanoma. Deregulations in cell signaling leading to increases in cell size has been described as one of the hallmarks of cancer (Hanahan and Weinberg, 2011). However, such increases in cell size have been hard to measure until now and issues with reproducibility, time consumption and level of expertise required for reliable assessments have limited its utility both in research and clinic. Modern user-friendly digital image analysis techniques offer an attractive solution to these problems, and for the first time we can now objectively quantitate multiple morphometry parameters in thousands of cells at a time.

In turn, changes to the size and shape of tumor cells are but a consequence of changes in the genotype, epigenetics and environmental factors. These changes can be tested with alternative methods, such as gene expression tests, next generation sequencing and chromosome analysis (Robertson *et al*., 2017; Onken *et al*., 2012). As found by Onken *et al*. the helix-loop-helix inhibitor *ID2* suppress the epithelial phenotype associated with an enlarged nucleus (Onken *et al*., 2006). Loss of *ID2* up-regulates the epithelial adhesion molecule E-cadherin, which in turn promotes the anchorage-independent cell growth required for metastasis. Consequently, we regard the morphometric characteristics investigated here as biomarkers, primarily for the cancer genotype.

Limitations of this study include a limited sample size. Substantial investments in digital scanning capacity is required before the method presented here can be used. The time consumption specified does not include preanalytical operations such as digital scanning and loading and unloading of glass slides. Small changes in the settings of the software’s cell detection function will influence the results greatly and even though measurements are automatized and the interobserver concordance is almost perfect as shown here, the definition of representative regions of interest requires at least a basic experience in ophthalmic pathology. This may reduce the generalizability of our method, the number of potential users and its application in everyday clinical routine. Further, as only one region of interest per tumor is defined, intratumor heterogeneity is not taken into account. As we have shown previously, there is significant variation in tumor characteristics including nBAP-1 expression in different subregions of UM (Stålhammar *et al*., 2019b). Last, our sample is not representative of all patients with uveal melanoma. We have only investigated the feasibility of digital morphometry in enucleated specimens without previous plaque brachytherapy. A large proportion of patients with uveal melanoma undergo primary plaque brachytherapy or proton beam radiotherapy and may never require enucleation. It remains unclear if the digital morphometry characteristics of small uveal melanomas is different from the relatively large tumors investigated here. Accordingly, we encourage future studies to confirm these results in larger cohorts that includes smaller tumors.

## Data Availability

The results published here are in part based upon data generated by the TCGA Research Network: https://www.cancer.gov/tcga.
All data analyzed during this study are included in this published article. The raw datasets generated during and/or analysed during the current study are available from the corresponding author on reasonable request.

## Acknowledgement

The results published here are in part based upon data generated by the TCGA Research Network: https://www.cancer.gov/tcga.

